# Pooled Multiplex Amplicon Massive Parallel Sequencing for High-Throughput Confirmation of Whole-Genome Sequencing Potentially Pathogenic Variants in Patients with Parkinsonism

**DOI:** 10.64898/2026.11.10.26367697

**Authors:** Radek Vodicka, Kristyna Kolarikova, Magdalena Soukup-Vodickova, Katerina Mensikova, Petr Kanovsky, Marketa Kolarikova, Barbora Majerova, Radek Vrtel, Martin Prochazka

## Abstract

Whole-genome sequencing identifies large numbers of genetic variants, although only a subset requires independent confirmation for clinical interpretation. Sanger sequencing remains the standard confirmation method but is impractical for large-scale variant verification. We developed and evaluated a strategy for variant confirmation based on multi-amplicon massively parallel sequencing of pooled DNA samples. Whole-genome sequencing data were obtained from 86 individuals, including patients with parkinsonism and controls. Variants were selected using phenotype-driven filtering, ClinVar classification, and pathogenicity assessment criteria. A total of 307 variants were targeted using a custom amplicon panel comprising 290 amplicons. DNA samples were normalized, pooled into 16 mixtures, and sequenced following automated library preparation. Variant presence and sequencing quality were assessed using standard bioinformatic tools. Of the 307 selected variants, 243 were successfully confirmed. Unconfirmed variants included sequencing artifacts (18), insufficiently covered targets (5), large insertions or deletions exceeding 12 base pairs (36), and variants located within homopolymeric regions (5). Results for selected variants were concordant with Sanger sequencing, and observed variant frequencies corresponded to expected frequencies in pooled samples. This approach provides a robust, rapid, and cost-effective method for high-throughput genetic variants confirming identified by whole-genome sequencing while requiring only limited DNA input. Despite complete concordance with Sanger sequencing, larger validation studies are needed before multi-amplicon massively parallel sequencing can replace routine confirmatory testing.

**Key Points:**

- Design and application of a high-throughput verification strategy for WGS variants with minimal DNA input requirements
- Reliable and independent confirmation of WGS findings
- Utilization of multi-amplicon massively parallel sequencing (MPS) in sample mixtures
- Detection of point mutations, small indels and duplications.

## Introduction

High-throughput molecular genetic sequencing analyses can be broadly categorized based on the extent of human genome coverage into targeted sequencing, whole exome sequencing (WES), and whole genome sequencing (WGS). Targeted sequencing focuses on specific genes or genomic regions—ranging from several to hundreds of genes—that are known to be associated with a particular disease. This approach typically yields hundreds to thousands of genetic variants per sample, among which only a limited number are clinically relevant.^1–3^ Whole exome sequencing expands the scope to all protein-coding regions of the genome, encompassing approximately 1–2% of the genome but capturing the majority of known disease-causing variants.^4–6^ WES can identify up to hundreds of thousands of variants per individual, with dozens potentially having clinical significance.

The most comprehensive approach is WGS, which covers the entire genomic DNA sequence, including coding and non-coding regions, regulatory elements, and structural variants.^3,7,8^ This approach provides a comprehensive amount of genetic information, detecting approximately 3–5 million single-nucleotide variants (SNVs) and small insertions/deletions (indels), as well as thousands of larger structural variants per individual.^9,10^

In our study, WGS was performed on 86 unique DNA samples collected from an isolated population in the Hornacko region (South Eastern Moravia, Czech Republic), a population characterized by a high and familial burden of Parkinsonism.^11^ Such isolated populations represent invaluable resources in genetic research due to reduced genetic heterogeneity and increased prevalence of founder mutations.^12,13^ Each sample generated approximately 5 million SNVs and indels, along with thousands of structural variants, dozens of which may have potential clinical impact. All of these variants may require orthogonal confirmation depending on laboratory policy, variant type and quality metrics. It should be ensured that the variants are real and not artifacts of the initial sequencing technique, especially in a clinical context where very high accuracy is crucial.

Currently, Sanger sequencing remains the gold standard for confirming variants identified by next-generation sequencing (NGS), especially single-nucleotide variants and small indels.^14–16^ However, this method is practical primarily when validating a small number of variants per sample. Applying Sanger sequencing to verify the vast number of variants from WGS across all 86 samples would be highly laborious, costly, and would consume considerable amounts of DNA and reagents.

### Effectiveness and Limitations of Orthogonal Confirmation

Large-scale studies show that Sanger sequencing confirms over 99.9% of NGS variants, however, routine Sanger confirmation adds cost and time with limited benefit, and best practices now suggest restricting confirmation to select cases.^17–20^ Machine learning and quality metrics can reliably identify which NGS calls require confirmation.^20,21^

### Alternative and Complementary Approaches

Other orthogonal methods, such as cross-platform NGS, copy number variant pipelines, and barcode-free error validation, have been validated and can further reduce the need for traditional confirmation.^18,22–24^ Cross-platform benchmarking and machine learning models can achieve high accuracy and efficiency.^21,22^

Alternative validation methods, including orthogonal NGS techniques or digital PCR, may be more feasible for large-scale validation of WGS data.^25^

If you originally sequenced on a short-read technology platform, validation may be performed on another platform, such as PacBio or Oxford Nanopore (long-read technologies).^20^

Using a smaller gene panel that focuses on suspicious variant regions can also be considered orthogonal to whole-genome sequencing.^26^

To confirm the whole-genome or exome sequencing results, you can also focus on specific gene regions and variants using multiamplicon NGS sequencing.^22^

### The aims of this study

1. To design and apply a strategy for the verification of WGS variants that is not demanding on the initial amount of DNA and can, within the same sequencing run, reliably, independently and within the available time verify WGS findings. This strategy is based on multi-amplicon massively parallel sequencing of mixed samples. The amplicon sequencing strategy should be predominantly focused on the detection of point mutations, small deletions and insertions (indels) up to 12 bp
2. To use the Sanger sequencing approach for NGS result validation and for resolving ambiguous results (low coverage, homopolymeric or repetitive regions or insertion/deletion larger than 12 bp)
3. For confirmed variants, the expected allele ratios were compared with the observed ratios obtained from the sequencing data.

## Material and Methods

The study was approved by the Ethics Committee of Palacky University and University Hospital Olomouc, Czechia. Written informed consent was obtained from all participants after they were thoroughly informed about the study.

The patient cohort included:

21 patients from the Hornacko region

44 controls from the Hornacko region

21 polysymptomatic patients outside the Hornacko region

This study does not address the clinical impact of the identified variants. Therefore, given the objectives of the study, detailed demographic and clinical characteristics of the entire cohort are not provided by the authors.

### DNA isolation

Peripheral blood (9 ml), collected in EDTA tubes, served as the primary biological material for DNA isolation. DNA was isolated from 74 whole blood samples using the salting-out method^27^, and from 12 whole blood samples using the magnetic bead-based extraction (MagNA Pure, Roche Diagnostics). Samples obtained using both extraction procedures were subsequently normalized to the same DNA concentration of 10 ng/µL and pooled irrespective of the extraction method.

### Selection of variants from WGS data

Whole genome sequencing analyses were performed using the Illumina TruSeq DNA PCR-free library preparation kit (Illumina, San Diego, CA, USA) and sequenced on the NovaSeq 6000 platform (Illumina, San Diego, CA, USA). Data analysis and variant annotation were conducted using Varsome Clinical 13.1.6 (Saphetor SA, Lausanne, Switzerland). In each analyzed sample, approximately 5 million variants were identified, with about 200,000 located in exonic and splicing regions.

The selection of variants for the confirmation was based on two primary filtering strategies using Varsome Clinical.

The first filter focused on genes and variants associated with Parkinsonism, including the following phenotypes: Parkinsonism (HP:0001300), Parkinsonism with favorable response to dopaminergic medication (HP:0002548), and Juvenile-onset Parkinson’s disease (MONDO:0000828). This filter included 178 genes (Supplemental Table S1) selected according to VarSome Clinical version 13.1.2.^28^ Variants were further filtered using a population allele frequency threshold of <0.001 (gnomAD v3.1.2) and ACMG criteria with the classification as pathogenic, likely pathogenic and variant of uncertain significance (VUS).

The second filter was based on ClinVar classification, selecting only variants annotated as pathogenic or likely pathogenic (to the date 11.10.2025).

### Amplicon design

After applying both filters, a total of 307 variants (including 14 from mitochondrial (mt) DNA) were selected for confirmation (138 SNVs, 126 small indels and 43 larger indels ≥ 12). The primers for independent confirmation were successfully designed using Ion AmpliSeq™ Custom Panels via the AmpliSeq Designer tool (Thermo Fisher Scientific, Waltham, MA, USA). In total, 290 unique amplicons were used to evaluate the selected 307 variants. All the necessary information about the variants is included in Supplemental Table S2.

### DNA mixture preparation

Eighty-six DNA samples were first normalized to a uniform concentration of 10 ng/µL using Qubit (Thermo Fisher Scientific, Waltham, MA, USA) and then pooled into 16 DNA mixtures. The first 10 mixtures each contained DNA from 5 individual samples, while the remaining 6 mixtures included DNA from 6 samples each. Each pooled mixture was further diluted in nuclease-free water to a final concentration of 0.7 ng/µL.

### Multiamplicon (maMPS) library preparation

Automated library preparation was performed in a single run using the Ion Chef™ Instrument (Thermo Fisher Scientific, Waltham, MA, USA) and the AmpliSeq™ Kit for Chef DL8 (Thermo Fisher Scientific, Waltham, MA, USA), following the manufacturer’s protocol (Ion AmpliSeq Library Preparation on the Ion Chef System, MAN0013432, Thermo Fisher Scientific, Waltham, MA, USA). All 16 resulting libraries were subsequently subjected to template preparation using the Ion 510™/520™/530™ Kit-Chef on the Ion Chef™ Instrument, according to the standard protocol (Ion 510™, Ion 520™, and Ion 530™ Kit-Chef, MAN0016854, Thermo Fisher Scientific, Waltham, MA, USA). Nineteen variants were purposively selected for orthogonal Sanger sequencing to represent different maMPS evaluation categories and technical conditions. The subset comprised nine variants classified within categories A1–A3, seven Rule B variants, and three category C variants and included different variant types, genomic locations, and sequencing coverage profiles. This subset was intended as an orthogonal methodological check rather than a comprehensive analytical validation of maMPS against Sanger sequencing.

### Ion Torrent sequencing

Massively parallel sequencing was performed using the Ion S5™ System (Thermo Fisher Scientific, Waltham, MA, USA) with Ion 510™, Ion 520™, and Ion 530™ chemistry. Template preparation and sequencing were conducted according to the manufacturer’s recommendations described in the Ion 510™ & Ion 520™ & Ion 530™ Kit–Chef, MAN0016854.

### Torrent Suite bioinformatic data processing

Data analysis, including quality control, signal calibration, alignment (to the GRCh38.p2.mask1 reference genome), base (variant) calling, amplicon coverage assessment, and overall sequencing quality metrics, was performed using Torrent Suite Software version 5.12.2. (Thermo Fisher Scientific, Waltham, MA, USA). The key analysis parameters are as follows (Table 1):

**Table 1.** Ion Torrent sequencing and Torrent Suite data analysis parameters. column1 – Individual arguments for bioinformatics steps, column 2 – source evaluation procedures.

| **Parameter** | **Setting / Command** |
|---|---|
| Chip Type | 530 |
| Beadfind args | justBeadFind --args-json /opt/ion/config/args_530_beadfind.json |
| Analysis args | Analysis --args-json /opt/ion/config/args_530_analysis.json |
| Pre-BaseCaller Args for calibration | BaseCaller --trim-qual-cutoff 15 --barcode-filter-minreads 10 --phasing-residual-filter=2.0 --wells-normalization on |
| Calibration Args | Calibration --num-calibration-regions 1,1 |
| BaseCaller args | BaseCaller --trim-qual-cutoff 15 --barcode-filter-minreads 10 --phasing-residual-filter=2.0 --num-unfiltered 1000 --barcode-filter-postpone 1 --qual-filter true --qual-filter-slope 0.040 --qual-filter-offset 1.0 --wells-normalization on |
| Alignment args | tmap mapall -q 50000 ... stage1 map4 |
| IonStats args | ionstats alignment |
| Thumbnail BeadFind args | justBeadFind --args-json /opt/ion/config/args_530_beadfind.json --thumbnail true |
| Thumbnail Analysis args | Analysis --args-json /opt/ion/config/args_530_analysis.json --thumbnail true |
| Thumbnail Pre-BaseCaller Args | BaseCaller --trim-qual-cutoff 15 --barcode-filter-minreads 10 --phasing-residual-filter=2.0 --wells-normalization on |
| Thumbnail Calibration Args | Calibration |
| Thumbnail BaseCaller args | BaseCaller --trim-qual-cutoff 15 --barcode-filter-minreads 10 --phasing-residual-filter=2.0 --qual-filter true --qual-filter-slope 0.040 --qual-filter-offset 1.0 --wells-normalization on |
| Thumbnail Alignment args | tmap mapall -q 50000 ... stage1 map4 |
| Thumbnail IonStats args | ionstats alignment |

### Setting of variant evaluation rules

For each variant, the expected read count ratio is determined in advance. The presence of variants was qualified and quantified using 16 BAM/BAI files for each mixture separately and the IGV Viewer (version 2.3.37). Variants are evaluated according to the following criteria:

A. **Variant classified as confirmed**

A1. The variant is considered confirmed if it is detected in the correct DNA mixture and the observed read ratio corresponds with the expected one.
A2. In cases of lower sequencing coverage ≤ 50 and/or if the observed ratio does not match the expected ratio (q ≤ 0.05). In such cases, a variant is considered as confirmed if it appears in the appropriate mixture and is absent from all the others.
A3. In homopolymer or repetitive regions where coverage is otherwise sufficient, the observed read count may still deviate from expected values. In such cases, we calculate the average number of false-positive reads in negative mixtures (where the variant was not identified by WGS) and subtract this background value from the observed read count in the relevant mixture. If the adjusted read count deviates from the expected value within a range corresponding to the variant’s mixture ratio, the variant is considered confirmed.
B. **Variant classified as unconfirmed (maMPS-unconfirmed/presumed WGS artifact)** The variant is not detected in the appropriate DNA mixture, and it is not located in a homopolymer region or involves insertion/deletion larger than 12 bp.
C. **Variant classified as non-evaluable**

C1. The sequencing coverage is too low (≤ 50 reads), and at same time, the variant is not present.
C2. The variant is in a homopolymer region where the signal appears uniformly across all mixtures, making it indistinguishable.
C3. The variant involves a large insertion or deletion that may not be reliably detected (discussed in more detail in the Discussion section).

### Variant evaluation using Sanger sequencing

The validation of our methodology was performed using Sanger sequencing for 19 selected variants (Table 2). These variants were chosen to represent a range of variant types, genomic locations, and read coverage profiles to ensure a robust and representative assessment of the accuracy of our approach.

**Table 2.**
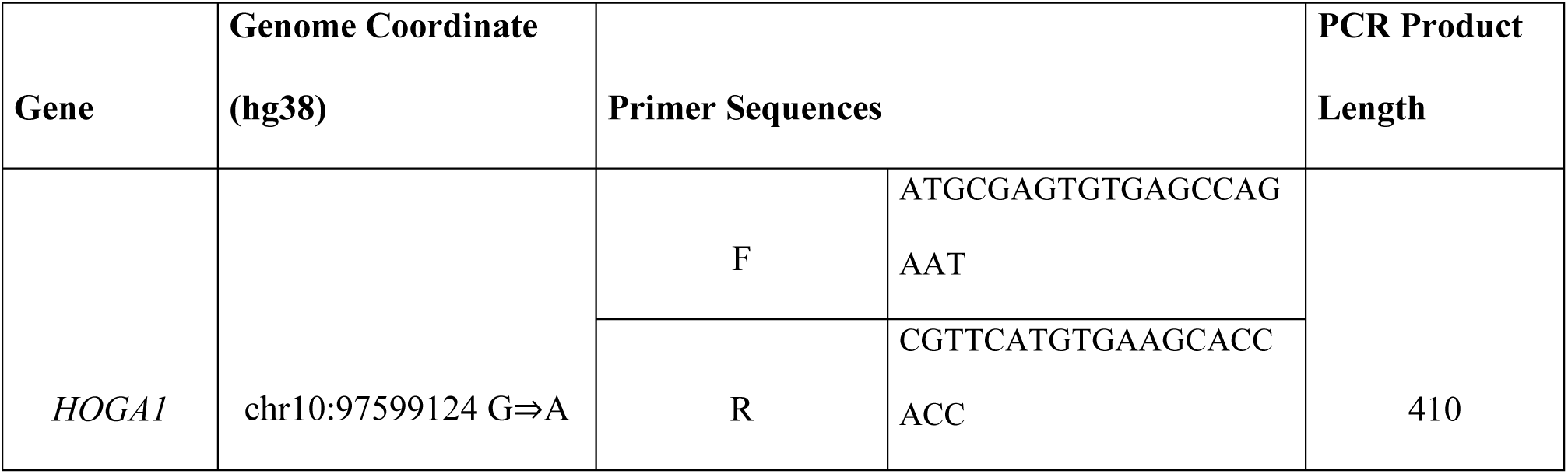
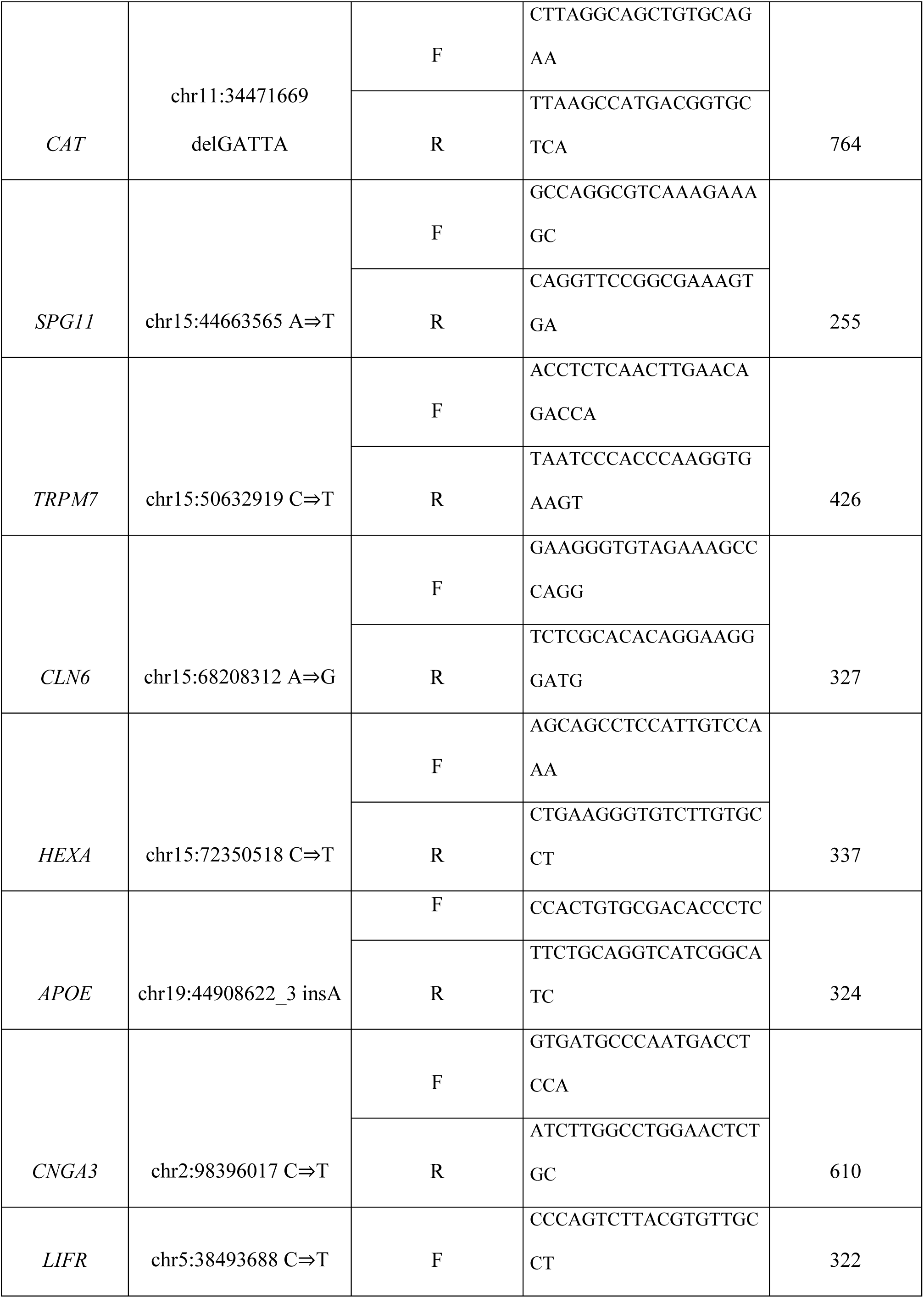
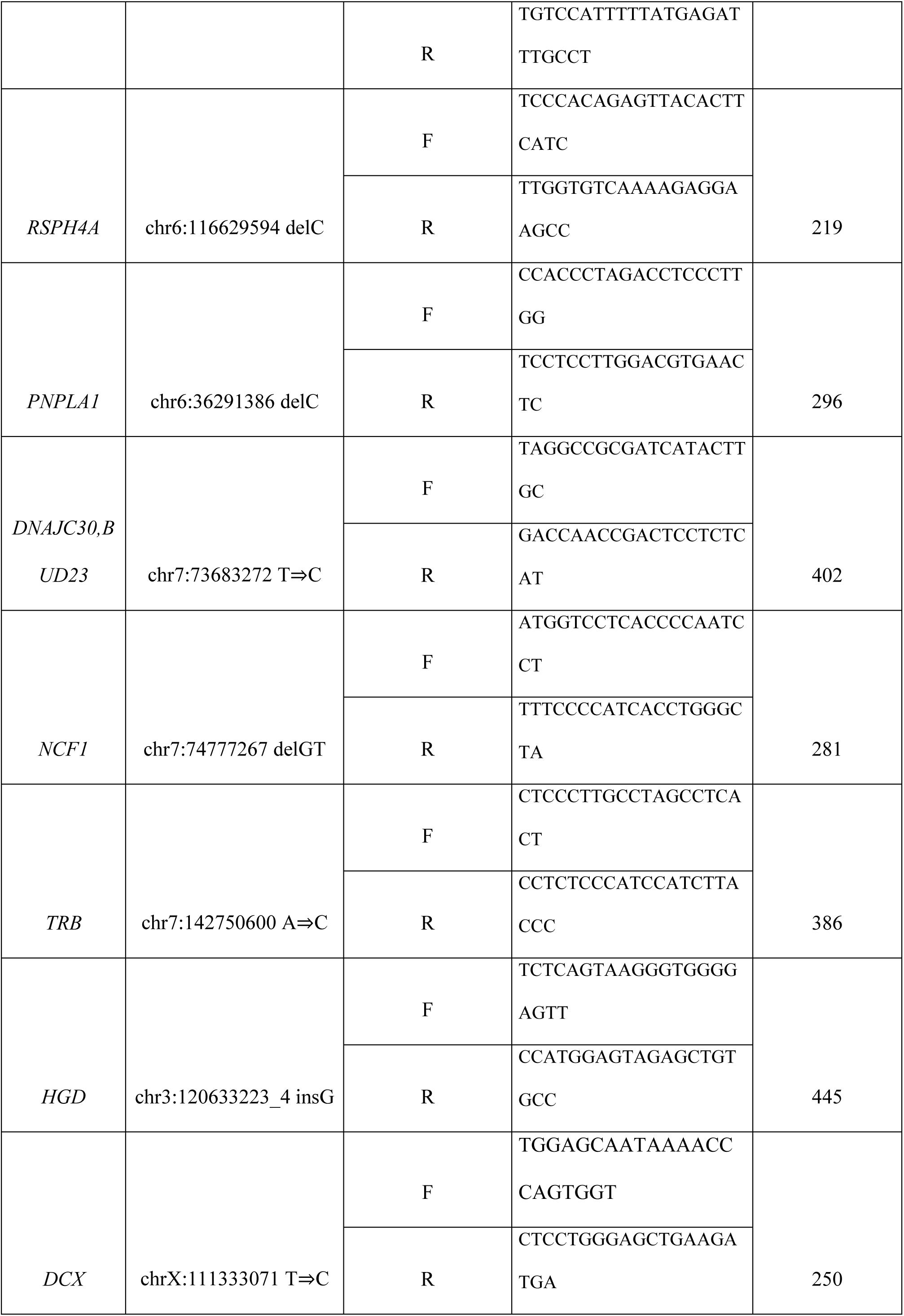
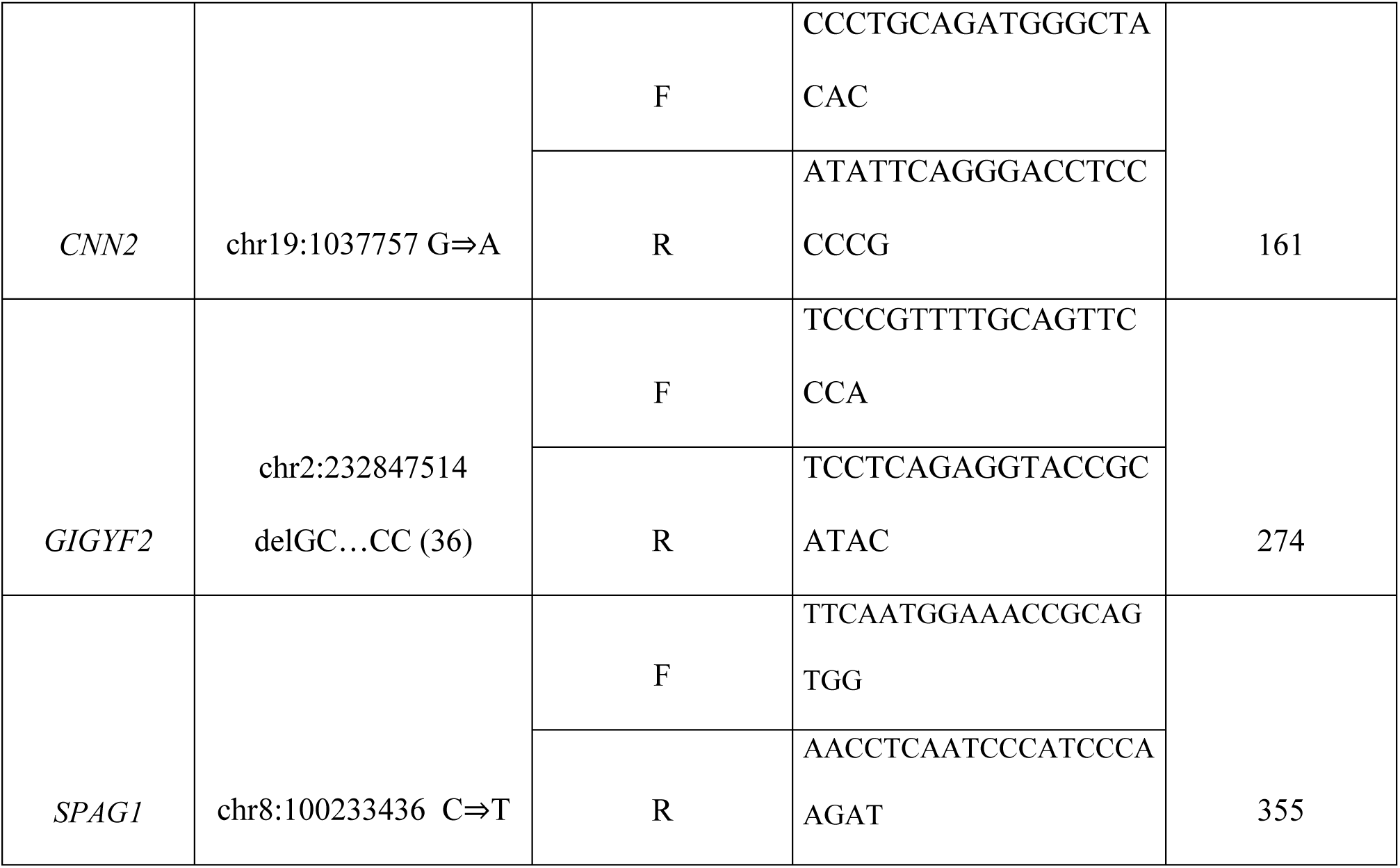
Selected primers for validation study. Gene refers to the specific gene in the human genome. Genomic region coordinates (GRCh38) indicate the chromosomal location based on the GRCh38 reference genome. Primer sequences are shown as F (forward) and R (reverse). PCR product length is indicated as size in base pairs. Primers used for Sanger sequencing were designed using Primer-BLAST web tool (National Center for Biotechnology Information, NCBI), and were not derived from previously published primer sets.

### Statistical expected and observed ratio comparison methods

For each mixed-DNA sample, we compared in A category (A3 homopolymeric and mitochondrial regions were not included) the observed number of variants (k) out of the total number of observations (n) to a theoretical expected proportion (p0) derived from the mixture design. Deviations from the expected proportion were assessed using an exact two-sided binomial test (H0: p = p0). To account for multiple testing across samples, p-values were adjusted using the Benjamini–Hochberg procedure to control the false discovery rate (FDR). Analyses were performed in R version 4.5.2 (2025 10-31-ucrt) see Supplemental Table S3.^30^

## Results

The numbers of mapped reads and the average coverage for each mixture are summarized in Table 3.

**Table 3.** Number of mapped reads and average coverage. Barcode ID: Unique identifier assigned to each sequencing library; Sample Name: Project and sample mixture name; Mapped Reads: Total number of sequencing reads successfully aligned to the reference genome; On Target: Percentage of reads mapping to the intended target regions; Mean Depth: Average sequencing coverage depth over the target regions.

| **Barcode ID** | **Sample Name** | **Mapped Reads** | **On Target** | **Mean Depth** |
|---|---|---|---|---|
| IonCode_0101 | WGS _confirm_Sample 1-5 | 191428 | 91.11% | 578.0 |
| IonCode_0102 | WGS _confirm_Sample 6-10 | 224866 | 89.94% | 664.2 |
| IonCode_0103 | WGS _confirm_Sample 11-15 | 265137 | 90.19% | 795.4 |
| IonCode_0104 | WGS _confirm_Sample 16-20 | 186509 | 91.26% | 567.3 |
| IonCode_0105 | WGS _confirm_Sample 21-25 | 174794 | 91.04% | 523.3 |
| IonCode_0106 | WGS _confirm_Sample 26-30 | 235473 | 90.54% | 710.8 |
| IonCode_0107 | WGS _confirm_Sample 31-35 | 214871 | 91.95% | 656.6 |
| IonCode_0108 | WGS _confirm_Sample 36-40 | 238878 | 93.08% | 742.2 |
| IonCode_0109 | WGS _confirm_Sample 41-45 | 244879 | 91.77% | 745.8 |
| IonCode_0110 | WGS _confirm_Sample 46-50 | 346727 | 92.04% | 1059 |
| IonCode_0111 | WGS _confirm_Sample 51-56 | 57438 | 91.22% | 171.7 |
| IonCode_0112 | WGS _confirm_Sample 57-62 | 241231 | 92.13% | 737.4 |
| IonCode_0113 | WGS _confirm_Sample 63-68 | 330969 | 92.50% | 1015 |
| IonCode_0114 | WGS _confirm_Sample 69-74 | 310813 | 92.31% | 949.7 |
| IonCode_0115 | WGS _confirm_Sample 75-80 | 343766 | 91.58% | 1048 |
| IonCode_0116 | WGS _confirm_Sample 81-86 | 271319 | 91.41% | 827.1 |

A total of 693 variants were assessed from 86 samples (the number of variants differs from the number designed by AmpliSeq designer (307) as many of the particular variants were found in more than one sample). The number of the variants evaluated according to the specified criteria is shown in Table 4.

**Table 4.** Summary of confirmed and unconfirmed variants by maMPS. Variants were classified according to predefined confirmation criteria. Rule A1 represents variants fulfilling the standard confirmation criteria; Rule A2 includes confirmed variants with sequencing coverage ≤50 and/or q ≤0.05. Rule A1/A2 indicates variants that fulfilled different confirmation criteria when detected in different sample mixtures. Rule A3 comprises confirmed variants located in homopolymeric regions. Unconfirmed/non-evaluable variants were classified as Rule B (maMPS-unconfirmed/presumed WGS artifacts), Rule C1 (insufficient or absent coverage), Rule C2 (variants in homopolymeric regions preventing reliable confirmation) and Rule C3 (indels ≥12 bp). Values are presented as the number of unique variants and the number of variant observations in separate columns.

| **Category and its rules** | **Number of unique variants** | **Number of variant observations** |
|---|---|---|
| **Confirmed variants (total)** | 243 | 347 |
| • Rule – A1 | 189 | 257 |
| • Rule – A2 (coverage ≤ 50 and/or q ≤ 0.05) | 24 | 34 |
| • Rule – A1/A2 (the same variant in different mixtures may meet both criteria) | 1 | 9 |
| • Rule – A3 (homopolymeric regions) | 29 | 47 |
| **Unconfirmed / non-evaluable variants (total)** | 64 | 346 |
| • Rule - B (sequencing artifacts) | 18 | 252 |
| • Rule - C1 (not covered) | 5 | 5 |
| • Rule - C2 (homopolymeric regions) | 5 | 5 |
| • Rule – C3 (indels ≥12) | 36 | 81 |

A total of 307 unique WGS-derived variants were included in the maMPS panel. Because several variants were present in more than one individual, this corresponded to 693 individual variant observations across the 86 samples. Overall, 243 of 307 unique variants were confirmed by maMPS. Of these, 189 variants fulfilled the standard A1 confirmation criteria, 24 fulfilled the A2 criteria, one variant fulfilled A1 or A2 depending on the individual mixture, and 29 variants were classified according to the A3 criteria for homopolymeric regions.

Among the remaining 64 unique variants, 18 were not detected by maMPS despite technically evaluable sequencing conditions and were classified as maMPS-unconfirmed/presumed WGS artifacts (Rule B). Five variants could not be assessed because of insufficient coverage (C1), five were non-evaluable because of homopolymeric sequence context (C2), and 36 represented indels ≥12 bp that could not be reliably assessed using the current amplicon design (C3). After excluding large indels ≥12 bp, 254 of 264 SNVs and small indels (96.2%) were technically evaluable using the proposed workflow.

Examples of assessments in IGV are shown in Figures 1 and 2.

**Figure 1:**
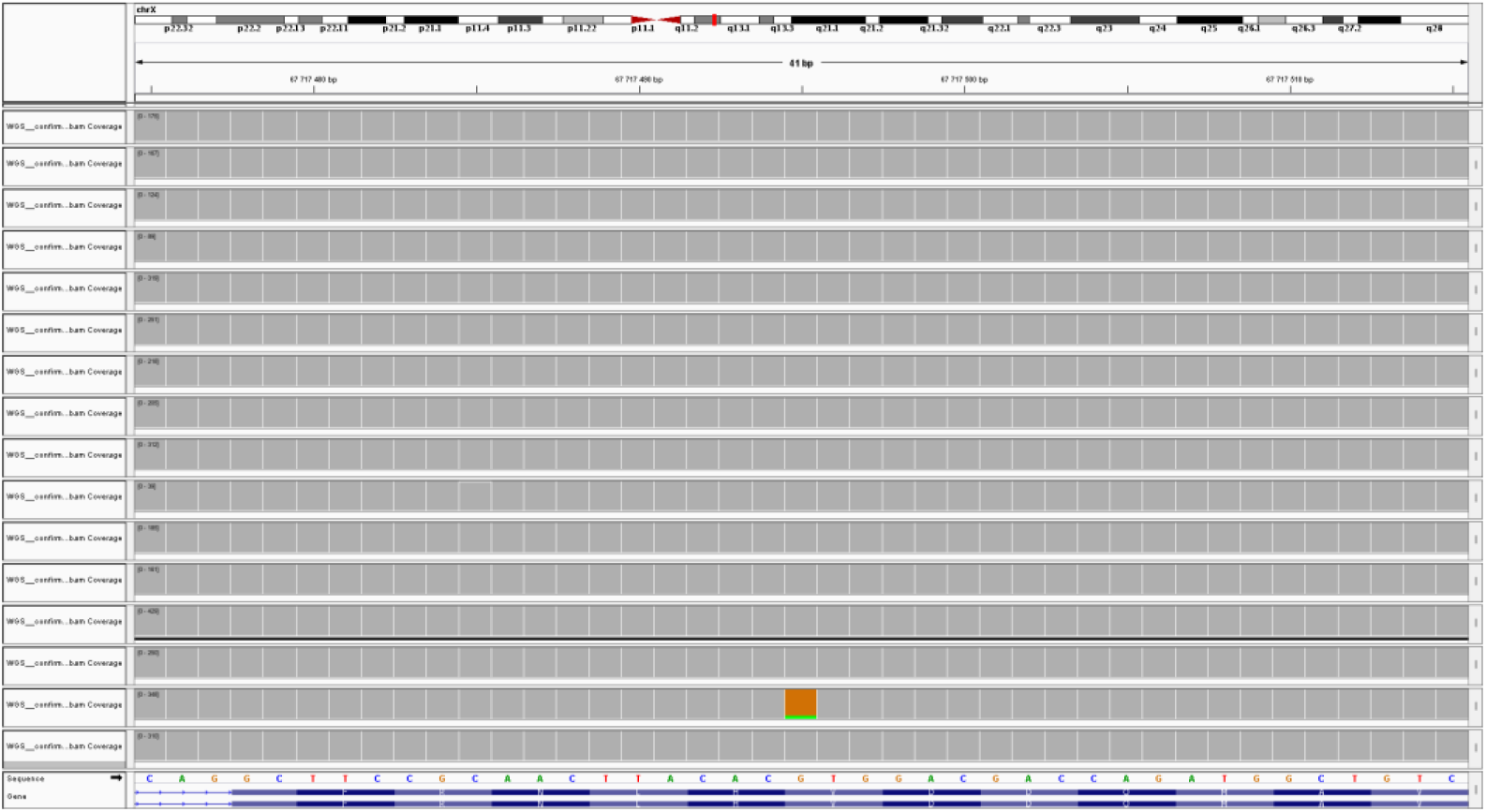
Confirmation of variant rs137852571 visualized using software IGV. In the left columns, individual mixtures are shown. Grey columns indicate sequencing coverage, and the reference genome (GRCh38) is visualized at the bottom. The confirmed variant rs137852571 (green bar) is shown in mixture 15 of sample 79. The expected A/G allele ratio of 0.083 is consistent with the observed ratio of 0.10 (number of A/G reads is 35/312).

**Figure 2:**
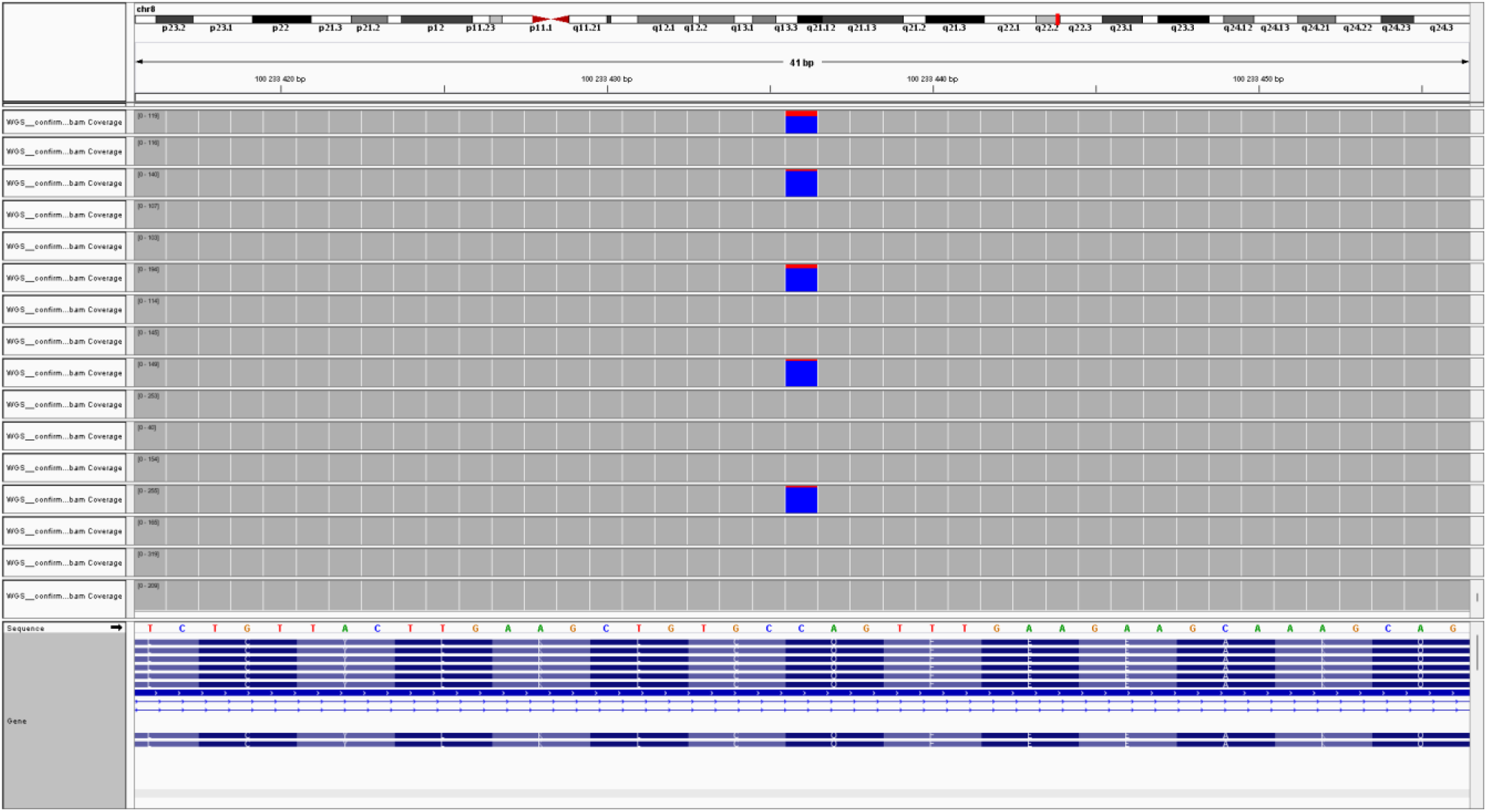
Confirmation of variant rs201740530 visualized using software IGV. In the left columns, individual mixtures are shown. Grey columns indicate sequencing coverage, and the reference genome (GRCh38) is visualized at the bottom. The confirmed variant rs201740530 (red bar) is shown in several mixtures. Mixture 1 in samples 3 and 4: expected T/C ratio 0.2 is consistent with the observed 0.2 (the number of T/C reads is 24/95); Mixture 3 in samples 14: expected T/C ratio 0.1 is consistent with the observed 0.09 (the number of T/C reads is 13/126); Mixture 6 in samples 26: expected T/C ratio 0.1 is not consistent with the observed 0.16 (the number of T/C reads 32/162); Mixture 9 in samples 42: expected T/C ratio 0.1 is consistent with the observed 0.09 (the number of T/C reads is 14/135); Mixture 13 in samples 65: expected T/C ratio 0.1 is consistent with the observed 0.08 (the number of T/C reads is 20/234)

Nineteen selected variants were additionally investigated by Sanger sequencing. All nine variants classified as confirmed by maMPS according to Rules A1–A3 were also confirmed by Sanger sequencing. Among seven Rule B variants, six were likewise not detected by Sanger sequencing, whereas one variant was confirmed by Sanger sequencing. Two low-coverage C1 variants that could not be evaluated by maMPS were detected by Sanger sequencing, while the C3 variant was not confirmed by either method. These results demonstrate good concordance for maMPS-confirmed variants while also illustrating the importance of orthogonal testing for technically challenging or non-evaluable findings. Table 5 provides an overview of particular variants according to the different rules that were evaluated using Sanger sequencing.

**Table 5.** Summary of confirmation analysis results. Genomic region coordinates (GRCh38) indicate chromosomal positions according to the GRCh38 reference genome. Gene denotes the corresponding human gene. WGS results represent findings from primary WGS data. MaMPS evaluation rules indicate the established criteria used for variant assessment. Sanger sequencing represents the results of confirmatory analysis.

| Genome region coordinates (GRCh38) | Gene | WGS result | maMPS | maMPS evaluation rule | Sanger sequencing |
|---|---|---|---|---|---|
| chr10:97599124 G⇒A | *HOGA1* | original finding | confirmed | A2 | confirmed |
| chr11:34471669 delGATTA | *CAT* | original finding | unconfirmed | B | confirmed |
| chr15:44663565 A⇒T | *SPG11* | original finding | confirmed | A2 | confirmed |
| chr15:50632919 C⇒T | *TRPM7* | original finding | unconfirmed | B | unconfirmed |
| chr15:68208312 A⇒G | *CLN6* | original finding | confirmed | A2 | confirmed |
| chr15:72350518 C⇒T | *HEXA* | original finding | confirmed | A2 | confirmed |
| chr19:44908622_3 insA | *APOE* | original finding | unconfirmed | B | unconfirmed |
| chr2:98396017 C⇒T | *CNGA3* | original finding | unconfirmed | C1 | confirmed |
| chr5:38493688 C⇒T | *LIFR* | original finding | unconfirmed | C1 | confirmed |
| chr6:116629594 delC | *RSPH4A* | original finding | confirmed | A3 | confirmed |
| chr6:36291386 delC | *PNPLA1* | original finding | confirmed | A3 | confirmed |
| chr7:73683272 T⇒C | *DNAJC30,*<br>*BUD23* | original finding | confirmed | A1 | confirmed |
| chr19:1037757 G⇒A | *CNN2* | original finding | unconfirmed | B | unconfirmed |
| chr7:74777267 delGT | *NCF1* | original finding | unconfirmed | B | unconfirmed |
| chr7:142750600 A⇒C | *TRB* | original finding | unconfirmed | B | unconfirmed |
| chr3:120633223_4 insG | *HGD* | original finding | confirmed | A3 | confirmed |
| chrX:111333071 T⇒C | *DCX* | original finding | unconfirmed | B | unconfirmed |
| chr2:232847514 delGC…CC<br>(36) | *GIGYF2* | original finding | unconfirmed | C3 | unconfirmed |
| chr8:100233436 C⇒T | *SPAG1* | original finding | confirmed | A1 | confirmed |

An example of Sanger confirmation is shown in the electrophoregram in Figure 3

**Figure 3:**
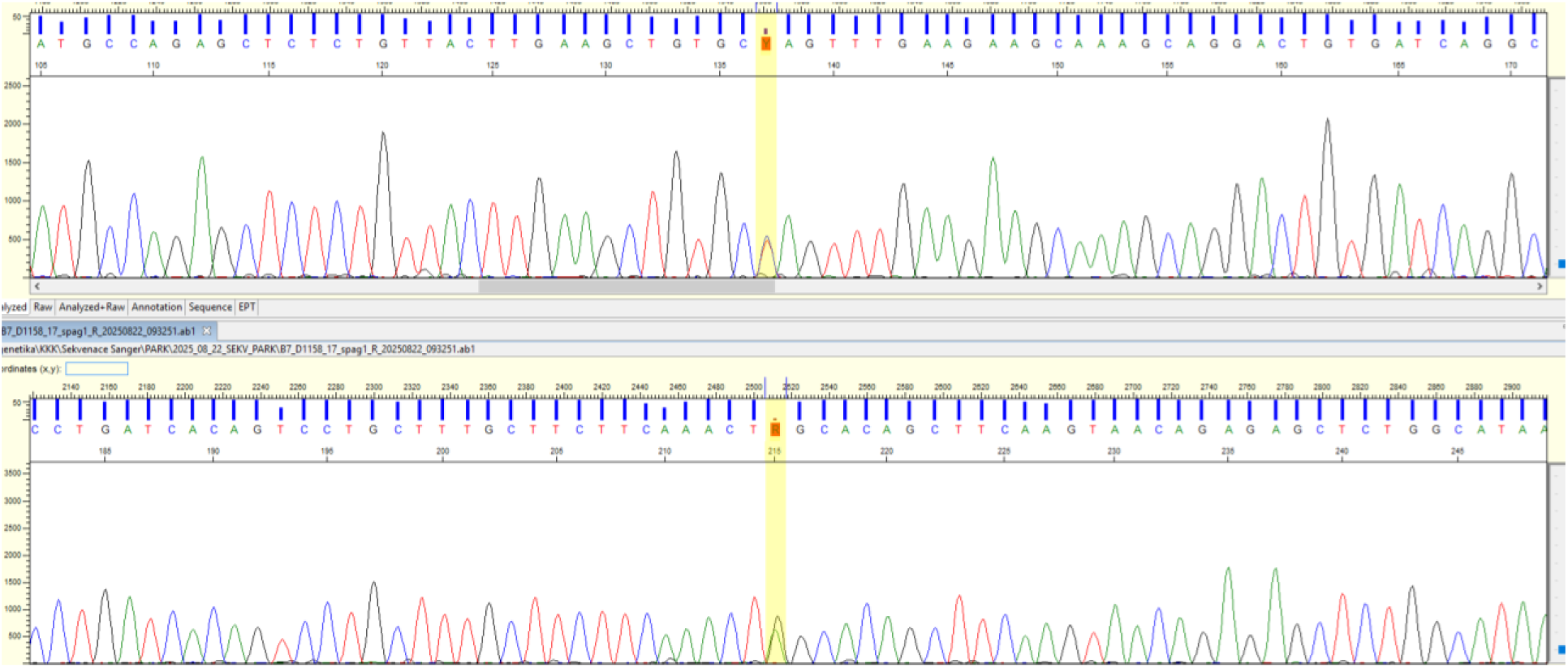
Confirmation of the rs201740530 (chr8:100233436 C⇒T) variant by Sanger sequencing. Representative bidirectional Sanger sequencing chromatograms showing the heterozygous rs201740530 variant. The variant position is highlighted in yellow. The forward sequence (upper panel) shows overlapping C/T peaks, while the reverse sequence (lower panel) shows the corresponding complementary G/A peaks, confirming the presence of the heterozygous C>T substitution.

### Result of the expected versus observed ratios comparison is summarized in Figure 4

Out of 278 (Supplemental Table 2) analyzed and confirmed variants (according A rule without homopolymeric and mtDNA variants), 33 (11.8%) showed significant deviation from the theoretical expected variant proportion after Benjamini–Hochberg FDR correction (q < 0.05), while 246 samples (88.2%) were consistent with expectation. Among the significant results, 7 samples (21.2 % of significant; 2.5 % of all tests) exhibited higher-than-expected variant proportions, whereas 26 samples (78.6 % of significant; 9.3 % of all tests) showed lower-than-expected proportions.

**Figure 4:**
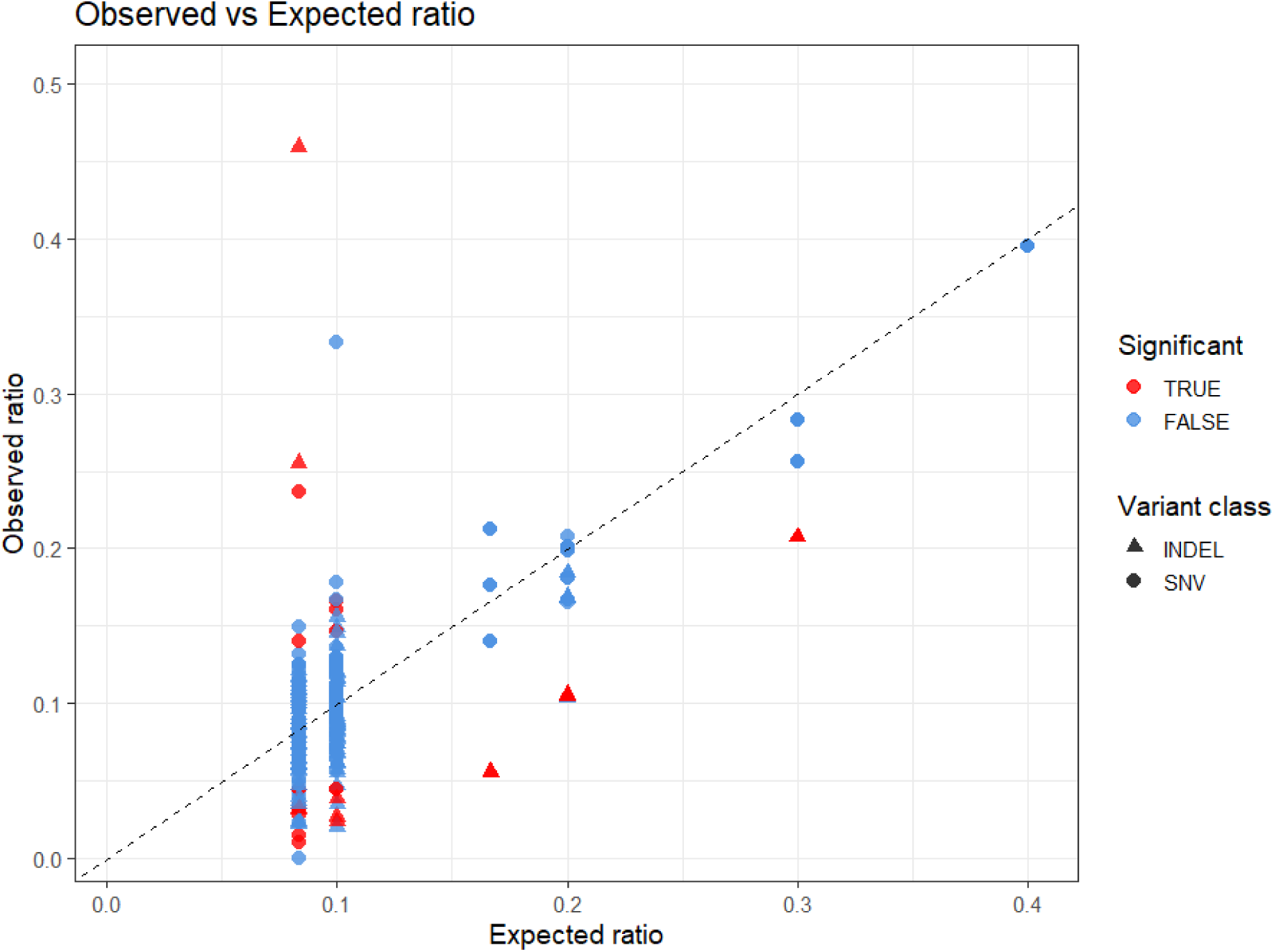
Scatter plot of observed vs expected variant allele frequency ratios. Scatter plot of observed (y-axis) versus expected (x-axis) variant allele frequency ratios for all samples. Each point represents an individual sample, with the dashed diagonal line indicating the line of perfect agreement between expected and observed ratios. Points closer to the diagonal reflect smaller deviations from expected proportions, whereas those farther from the line reflect greater deviations. Samples without significant deviation (q > 0.05) are shown in blue, while samples with significant deviation (q ≤ 0.05) are shown in red. Triangles correspond to insertions/deletions (indels), and circles correspond to single-nucleotide variants (SNVs).

Samples cluster in six discrete columns corresponding to the expected allele frequency ratios derived from pool composition (0.10, 0.20, 0.30 and 0.40 for five-sample mixtures and 0.083, 0.166 for six-sample mixtures). Most variants cluster closely along the diagonal, consistent with good agreement between observed and expected allele frequencies. Significant outliers deviate in both directions, reflecting bidirectional discordance between sequencing-derived and expected allele frequencies. Indels appear overrepresented among significantly deviating variants, sequencing method estimates the allele frequency of indels less accurately than that of SNVs.

## Discussion

NGS has become a common standard in clinical genetics testing. In clinical practice, NGS workflows are often optimized for high sensitivity, which may lead to an increased rate of false-positive results. Nevertheless, high accuracy and reliability in the detection of genetic variants are essential for clinical diagnosis.

### Evolving Standards and Guidelines

Multiple professional organizations now recommend that each laboratory develop its own policy for orthogonal confirmation, considering factors such as test content, sequencing platform, variant type, and quality metrics. Universal confirmation is no longer required; instead, confirmation should be targeted to variants with lower sequencing quality confidence or unexpected results.^19,25^ Laboratories are encouraged to document and regularly review their confirmation policies.^19,25^

Traditionally, Sanger sequencing has been considered the gold standard for orthogonal confirmation of NGS-detected variants, helping to reduce the risk of false positives (FPs).

However, Sanger sequencing is not without limitations; for example, allelic dropout can result in false negatives.^31–33^ Furthermore, designing primers is complicated in homopolymeric regions and in genomic regions with high sequence similarity, such as pseudogenes.^34^ Chennagiri *et al*. recommend combining two different sequencing technologies that utilize distinct principles of DNA selection and sequencing.^18^ This approach allows for independent confirmation of the majority (∼95%) of identified variants.

Some studies focus on reducing confirmatory analysis, as the associated time and financial costs can be critical in clinical settings.^20^ For single-nucleotide variants (SNVs) with high quality scores, confirmatory testing may be considered redundant.^21,35^

Sanger sequencing remains necessary, primarily for confirmation of variants with low quality scores and for detecting larger insertions or deletions (indels)^36,37^, although in some cases, confirmation of indels up to 68 bp can be omitted.^31^ There are also genomic regions that are challenging to sequence with high quality, such as pseudogenes and regions with segmental duplications. The risk of false positives in these regions varies and is not fully addressed in the cited literature.^31^

Moreover, particular attention should be given to somatic and mosaic variants. Although not all such variants require verification, they demand a different approach when setting reliability parameters.

For the detection and quantification of specific variants, especially in circulating tumor DNA, digital droplet PCR (ddPCR) has been recommended due to its high sensitivity.^38,39^ Additionally, mass spectrometry genotyping has been described as a useful method for confirming NGS results in cases where Sanger sequencing is limited, such as in homopolymeric regions.^40,41^

This study does not focus on the clinical impact of the identified variants but it evaluates the possibility and reliability of verifying variants detected by WGS using a newly proposed methodological approach. This approach is based on amplicon-based resequencing, using a custom amplicon design generated with the AmpliSeq Designer and sequenced on the Ion Torrent platform. Given our long-standing experience of assessing mixed samples and mosaicism using various methodologies (QF-PCR, minisequencing, real-time PCR, and droplet digital PCR)^42–46^, this similar strategy was selected as an appropriate verification method. The novelty of the present approach does not lie in amplicon-based sequencing itself, which is an established methodology, but in the design of the confirmation workflow. Individual DNA samples were combined into predefined mixtures, thereby reducing 86 individual samples to 16 sequencing libraries. Because the composition of each pool was known, an expected allele fraction could be calculated for each selected WGS variant. Confirmation therefore relied on three complementary features: detection using an independent sequencing platform, occurrence in the expected DNA mixture, and agreement with the theoretically expected allele ratio. This combination provides an additional verification layer while considerably reducing the number of individual sequencing libraries required.

Considering the limited amount of available and often unique DNA, the study design was optimized to allow the assessment of hundreds of variants presented in mixed samples in a heterozygous state with an expected allele ratio as low as 1/12, provided sufficient sequencing coverage was achieved. The study confirmed the well-known limitation of evaluating variants in homopolymeric or repetitive regions. These regions were evaluated using a correction for false-positive variant calls (see Methods – Variant evaluation). Overall, five variants located within homopolymeric regions could not be reliably assessed (rs1203776328, rs35978415, rs952840023, rs113262693 and rs902007551).

Another limiting aspect was the evaluation of indels larger than 12 bp. As we anticipated a higher degree of DNA fragmentation in a substantial proportion of samples stored in the DNA biobank for more than 10 years, we used a pipeline optimized for the verification of SNPs and small indels during amplicon custom design. Consequently, indels larger than 12 bp were frequently not detectable (36 out of 43 variants). Exceptions included seven variants (rs1278020632, rs553779459, rs111033253, rs1790142871, rs879074973, NM_020919.4:c.2417+2308_2417+2309insTCCAAAATATAG p.?, ENST00000394991.8:c.306+38698_306+38699insTATCCAGAAAGTTATTT, p.?,).

To improve the detection of larger indels, a more appropriate strategy would be to use amplicons of the largest possible size. In the case of the AmpliSeq Designer, amplicons of up to 375 bp could be considered. Alternatively, additional verification using conventional Sanger sequencing or long-read technology may be used.

All 14 mtDNA variants were reliably confirmed in all samples, including two with 12% and 13% heteroplasmy, as determined by WGS.

An important limitation of the present study is the relatively small number of variants subjected to independent Sanger sequencing. Although all maMPS-confirmed variants included in this subset were concordant with Sanger sequencing, the analysis of 19 variants does not provide sufficient evidence to conclude that maMPS can universally replace Sanger-based confirmation. Rather, the present results support its use as a scalable complementary confirmation strategy that may substantially reduce the number of individual Sanger analyses required. A larger systematic comparison with orthogonal sequencing would be necessary to establish analytical sensitivity and specificity and to define circumstances in which Sanger confirmation could be safely omitted. This question will be addressed in our forthcoming follow-up validation study, which will build directly upon the findings of the present work.

Of the 307 maMPS-assessed variants, 19 were selected based on the predefined criteria (A–C) for confirmatory Sanger analysis (Table 5). Variants classified by maMPS in category A demonstrated full concordance between the two methods.

Six variants classified according to rule B were generally concordant and were not confirmed by either maMPS or Sanger sequencing. A single exception was observed in *CAT*(NM_001752.4):c.236_240del p.? variant, which was confirmed by Sanger sequencing but not by maMPS. This discrepancy could be influenced by the design of the multiplex amplicon panel and the reduced amplification efficiency of the target locus may also contribute to the lack of confirmation by maMPS.

Three variants classified in category C included 2 low-coverage variants (C1), both of which were not confirmed by maMPS but were confirmed by Sanger sequencing indicating that insufficient sequencing depth primarily affected the performance of the maMPS-based confirmation. One variant classified according to rule C3 was not confirmed by either maMPS or Sanger sequencing.

In our expected versus observed analysis, p0 corresponds to a theoretical DNA mixing ratio and does not reflect biological variation. Therefore, a significant binomial test reflects a deviation from expected laboratory ratio and may be caused by several non-biological factors. Four major mechanisms are listed below.

#### a) Pipetting and quantification variability

DNA mixtures depend on accurate volumes and reliable concentration measurements. Pipetting introduces imprecision, and quantification adds additional error, especially at low DNA inputs.^47,48^ These inaccuracies may alter sample proportions.

#### b) Uneven coverage and locus dropout

NGS data exhibit highly heterogeneous read depth across genomic regions.^49^ Low coverage increases random sampling error, while preferential amplification introduces systematic allelic bias, causing the observed allele fraction (k/n) to deviate from its expected value.^50^

#### c) Systematic bias in variant detection

Variant callers show locus-specific sensitivity influenced by GC content, sequence complexity, and alignment artefacts.^51^ These biases can lead to reproducible over- or under-calling of variants.

#### d) Overdispersion relative to the binomial model

The exact binomial test assumes that variance is fully determined by n and p0. Sequencing data, however, exhibit technical heterogeneity.^52^ Ignoring overdispersion can underestimate uncertainty and increase false positives. Beta-binomial models are therefore preferred for NGS data.^53,54^ Considering these factors, the significant deviations observed here are best interpreted as indicators of technical variability. To better distinguish true mixture inaccuracies from stochastic sequencing noise, future studies could employ beta-binomial frameworks, use technical replicates, or perform locus-specific normalization.

## Conclusion

After excluding the indel ≥12 bp variants that were deemed unreliable or non-evaluable by this method, we were able to accurately and reliably assess 254 out of 264 (over 96 %) of the SNPs and small indels. Our approach demonstrates robust performance, and with the incorporation of longer reads it could also be extended to verify larger genomic alterations, thereby increasing its applicability in genetic research and diagnostics of Parkinsonism and similar polygenic disorders. Although maMPS showed complete concordance with Sanger sequencing in the tested subset, larger validation studies are required before it can be considered a replacement for routine Sanger-based confirmation.

## Data Availability

All data produced in the present study are available upon reasonable request to the authors.

## Author Contributions

R.V., K.M. and P.K. designed the study, K.K., M.S-V. and B.M. analyzed the data, R.V. contributed to data analysis and interpretation, R.V., K.K. M.S.-V. and M.K. drafted the manuscript. R.V. is the guarantor of this work and, as such, had full access to all of the data in the study and takes responsibility for the integrity of the data and the accuracy of the data analysis. all authors critically revised the manuscript, approved the final version, and agree to be accountable for all aspects of the work.

## Declaration of generative AI and AI-assisted technologies in the manuscript preparation process

During the preparation of this work, the authors used DeepL AI and ChatGPT for language editing and the formal formatting of tables and figures. The authors reviewed and edited the output as needed and take full responsibility for the content of the published article.

